# Impacts of school closure due to COVID-19 on the mobility trend of Japanese citizens

**DOI:** 10.1101/2021.06.30.21259816

**Authors:** Nguyen Hai Nam, Anh PN. Nguyen, Bao-Tran Do Le, Abdelrahman Gamil Gad, Abdelrahman Sherif Mohamed Abdelnaeim Abdalla, Adnan Safi, Anh Gia Pham, Vo Duc Khanh, Nguyen Tien Huy

## Abstract

School closure was the only main control measure that Japan took into action from late February to late March in 2020. Accurate evaluation of how Japanese citizens responded to the impact of school closure remains a challenge. Data from the Google COVID-19 Community Mobility Report was used to analyze the mobility trend of Japanese citizens regarding six categories, including retail and recreation, grocery and pharmacy, parks, transit stations, workplace, and residential. The median percentage of mobility in all 47 prefectures of Japan was calculated during five periods of time, including one week before school closure, one week, two weeks, three weeks, and four weeks after school closure. There was a significant decline in the mobility trend of transit stations, grocery and pharmacy, parks, retail and recreation, and workplace at the moment after school closure compared to the prior period. Inversely, the mobility trend in staying at home remarkably increased following the implementation of school closure. Our study determined a significant change in the mobility trend of Japanese citizens before and after school closure. These data reflected the responsibility and the consciousness of Japanese citizens in mitigating COVID-19.

## Introduction

Coronavirus is a large family of respiratory viruses that includes hundreds types of viruses^1^. A novel type of coronavirus, called severe acute respiratory syndrome coronavirus 2 (SARS-CoV-2), emerged in China in December 2019 and has caused coronavirus disease (COVID-19) worldwide^1^. On March 11^th^, 2020, the World Health Organization (WHO) declared the COVID-19 outbreak as a pandemic with 118,319 confirmed cases globally^2^. SARS-CoV-2 has a higher transmission rate than severe acute respiratory syndrome coronavirus (SARS-CoV) and middle east respiratory syndrome coronavirus (MERS-CoV), which are the other two coronaviruses causing previous epidemics^3^. Importantly, people mainly got COVID-19 by coming into close contact with another infected person^4^. Therefore, limiting social contacts is considered as a critical strategy to reduce the number of new COVID-19 cases.

Different control measures have been applied to control and mitigate the outbreak of COVID-19 depending on the specific situation and policies of each country, such as locking down affected areas, imposing entry restrictions, canceling mass gathering events, and closing schools and workplaces. In the Community Mitigation Guidelines to Prevent Pandemic Influenza published by the Centers for Disease Control and Prevention (CDC) in April 2017, school closure was among one of the most critical strategies to maintain social distancing, thus helping mitigate the pandemics^5^. Since the transmission of influenza and COVID-19 share many similarities^6,7^, several guidelines to control influenza have been applied for COVID-19, including school closure^8^. As of March 18^th^, 2020, 107 countries had canceled schools to control the COVID-19 pandemic^9^. Regarding Japan, from the end of February to the end of March, the main control measure that Japan took into action was to close schools nationwide. School closure started on March 2^nd^ to limit the spread of the virus transmission when the country had 241 confirmed cases^10^. Specifically, elementary, junior high, and high schools were ordered to close until early April^11^.

Accurate evaluation of how Japanese citizens responded to the impact of school closure remains a challenge. Evidence of the effects of school closures on the behavioral changes of individuals over time is scarce. Since the individual data of Japanese citizens are confidential and highly protected, the COVID-19 Community Mobility Report from Google, which is based on the location history of GPS of users, remains an accessible and reliable database. Thus, in this paper, we evaluated how school closure impacted the mobility of people living in Japan by using data from the COVID-19 Community Mobility Report of Japanese citizens.

## Methods

### Database

Google provides the Google COVID-19 Community Mobility Reports by using historical GPS data of users around the world^12^. Data from this report presented the percentage change of the mobility of Japanese citizens at several specific places, including retail and recreation, grocery and pharmacy, parks, transit stations, workplace, and residential.

These six categories recorded in the Google COVID-19 Community Mobility Reports were defined as (1) retail and recreation: the mobility trend in closed spaces such as restaurants, coffee shops, department stores, museums, libraries, and movie theaters; (2) grocery and pharmacy: the mobility trend in locations such as supermarkets, convenience stores, markets, and drugstores; (3) parks: the mobility trend in public areas such as parks, beaches, and harbours; (4) transit stations: the mobility trend in public transportation such as subways, railways, and buses; (5) workplace: the mobility trend in going to workplaces; and (6) residential: the mobility trend in staying at home.

### Data analysis

We compared the percentage of mobility in six categories during five periods of time, including one week before school closure (from February 24^th^ to March 1^st^), one week after school closure (from March 2^nd^ to March 8^th^), two weeks after school closure (from March 2^nd^ to March 15^th^), three weeks after school closure (from March 2^nd^ to March 22^th^), and four weeks after school closure (from March 2^nd^ to March 29^th^). Since the mobility report was a line graph, we used WebPlotDigitizer software to extract the percentage of these six categories in all 47 prefectures of Japan (https://automeris.io/WebPlotDigitizer/). The data obtained from the software was the percentage of the daily mobility of each prefecture. We then calculated the mobility percentage of a particular week by averaging the mobility percentage of all days during that period of time. After having the average mobility data in five periods of time of each prefecture, we calculated the median of mobility in all 47 prefectures to have the national data. A box and whisker plot was plotted to interpret the changes of mobility in Japan from February 24^th^ to March 29^th^.

### Statistical analysis

Data were tested for deviations from a normal distribution within the groups using a Shapiro-Wilks test. The percentage of mobility in each category was expressed as the median with interquartile and was compared between each other using the Mann–Whitney test. Statistical significance was set at p < 0.05. All statistical analyses were performed by JMP version 12 (SAS Institute, Inc., Cary, NC, USA).

## Results

The mobility of people living in Japan for retail and recreation witnessed a reduction after schools started closing nationwide. Compared to one week before school closure, the median (interquartile) mobility of retail and recreation significantly decreased at one week, two weeks, three weeks, and four weeks after school closure (1.6%(−4%;3.6%) vs. -8%(−10%;-5%); -6%(−7.8%;-3%); -3.8%(−6%;-1%); -3.8%(−5.7%;-2.7%). The lowest mobility percentage in this category was in period C (one week after school closure) with a reduction of 9.6% compared to one week before school closure. After the remarkable decline in period C, there was a trend to restore the mobility of retail and recreation, illustrated by the elevation in percentage of mobility in period D, E, and F (Figure 1).

**Figure 1:**
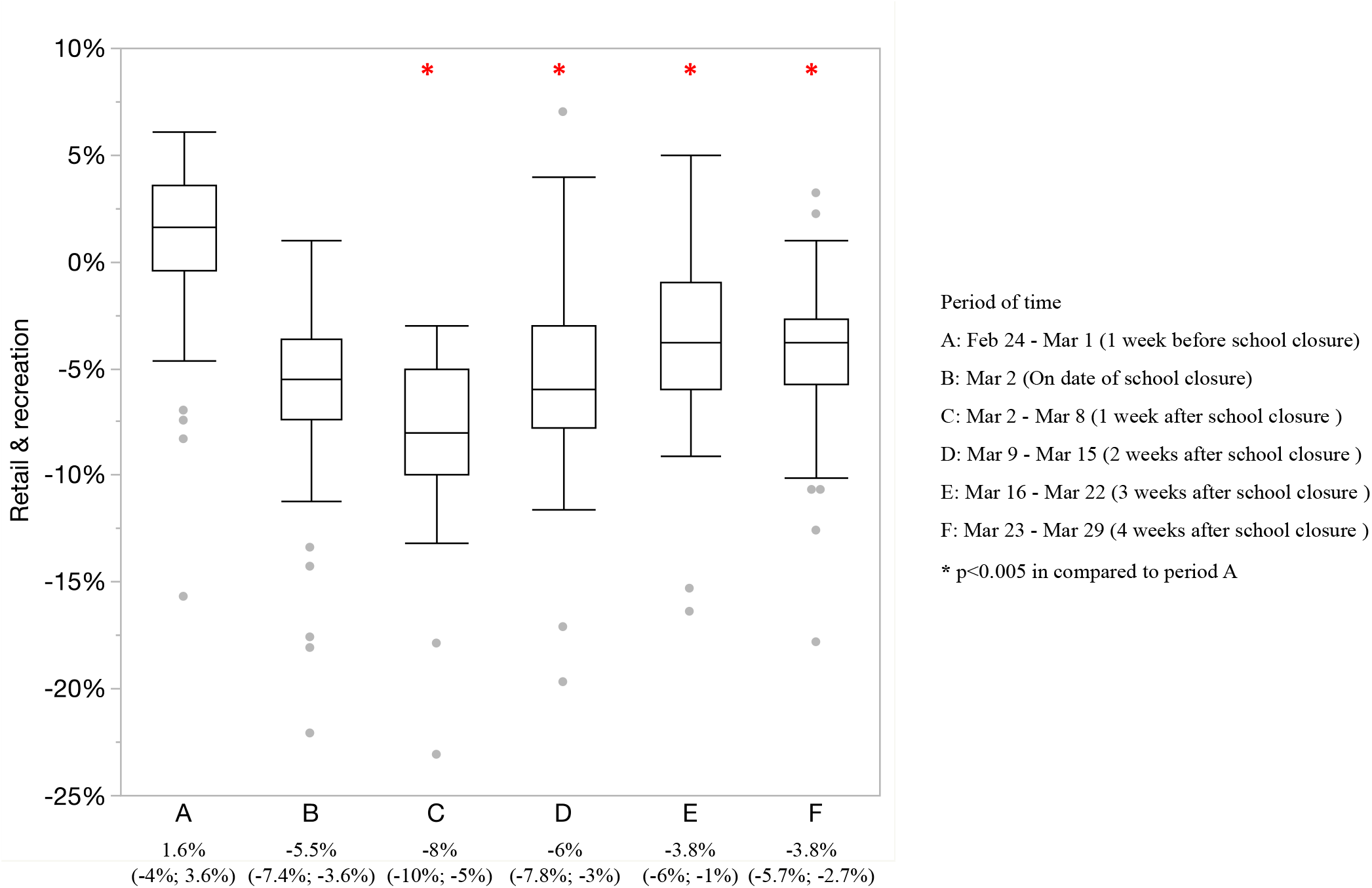
Mobility trend in retail and recreation of Japanese citizens from February 24^th^ to March 29^th^.

In a similar manner as retail and recreation, the median (interquartile) mobility of Japanese citizens regarding grocery and pharmacy significantly decreased at period C, D, E, and F compared to period A (1.7%(0.4%;2.3%); 1.7%(0.7%;2.4%); 2%(1%;3%), and 2.5%(1.5%;3.7%) vs. 7.4%(5.6%;8.2%)). Period C and D had the lowest percentage mobility of this category. A slight increase was identified after school closure since the moment of period D (Figure 2).

**Figure 2:**
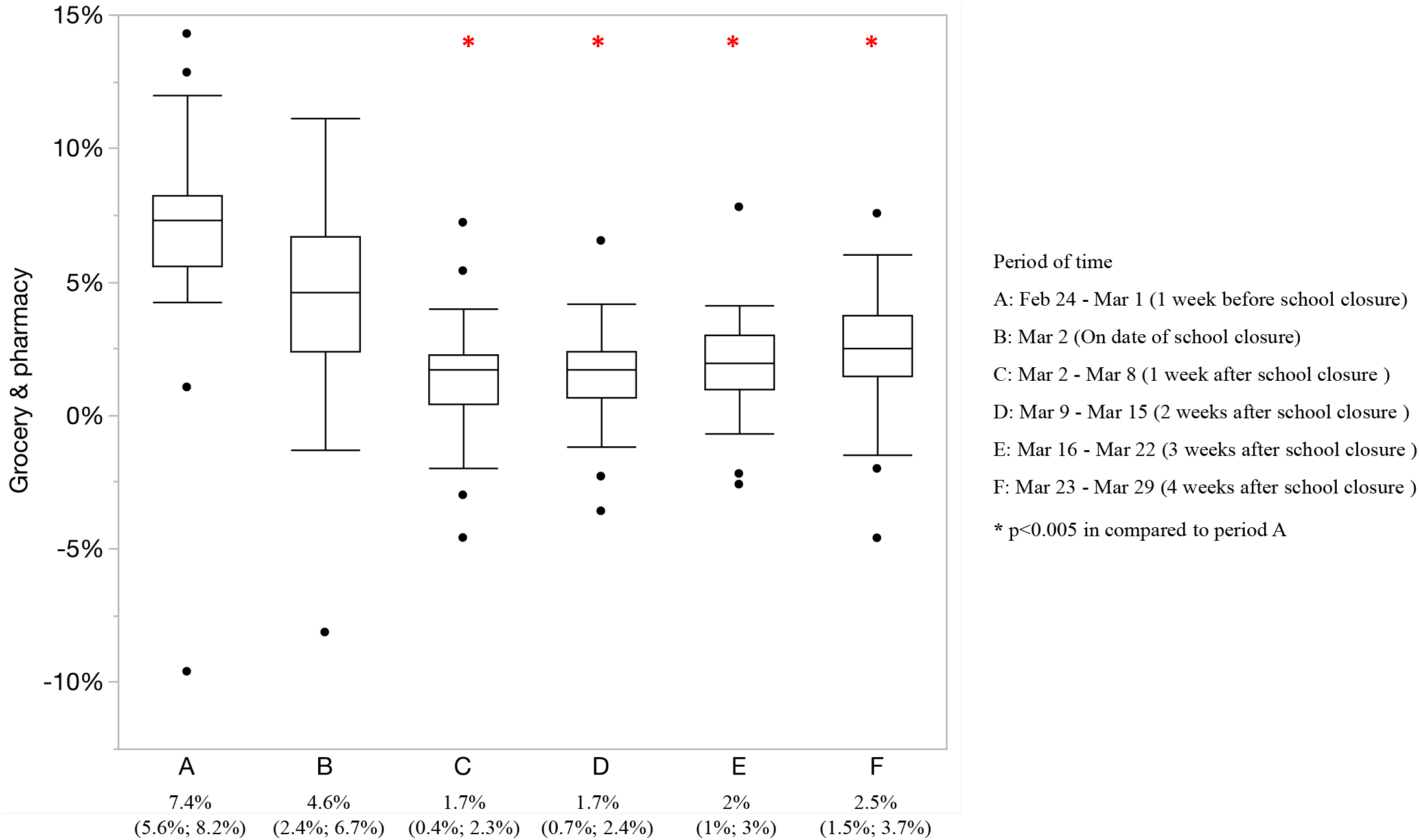
Mobility trend in grocery and pharmacy of Japanese citizens from February 24^th^ to March 29^th^.

Regarding the mobility trend for parks in Japan, a notable depletion of the median (interquartile) mobility was observed at period C and D compared to period A (−6%(−11.5%;-3%); -3.2(−8.8%;-1%) vs. 4.6%(1.6%;5.3%)). However, at the moment of three and four weeks after school closure, the mobility trend for parks restored as similar to that of one week before school closure (Figure 3).

**Figure 3:**
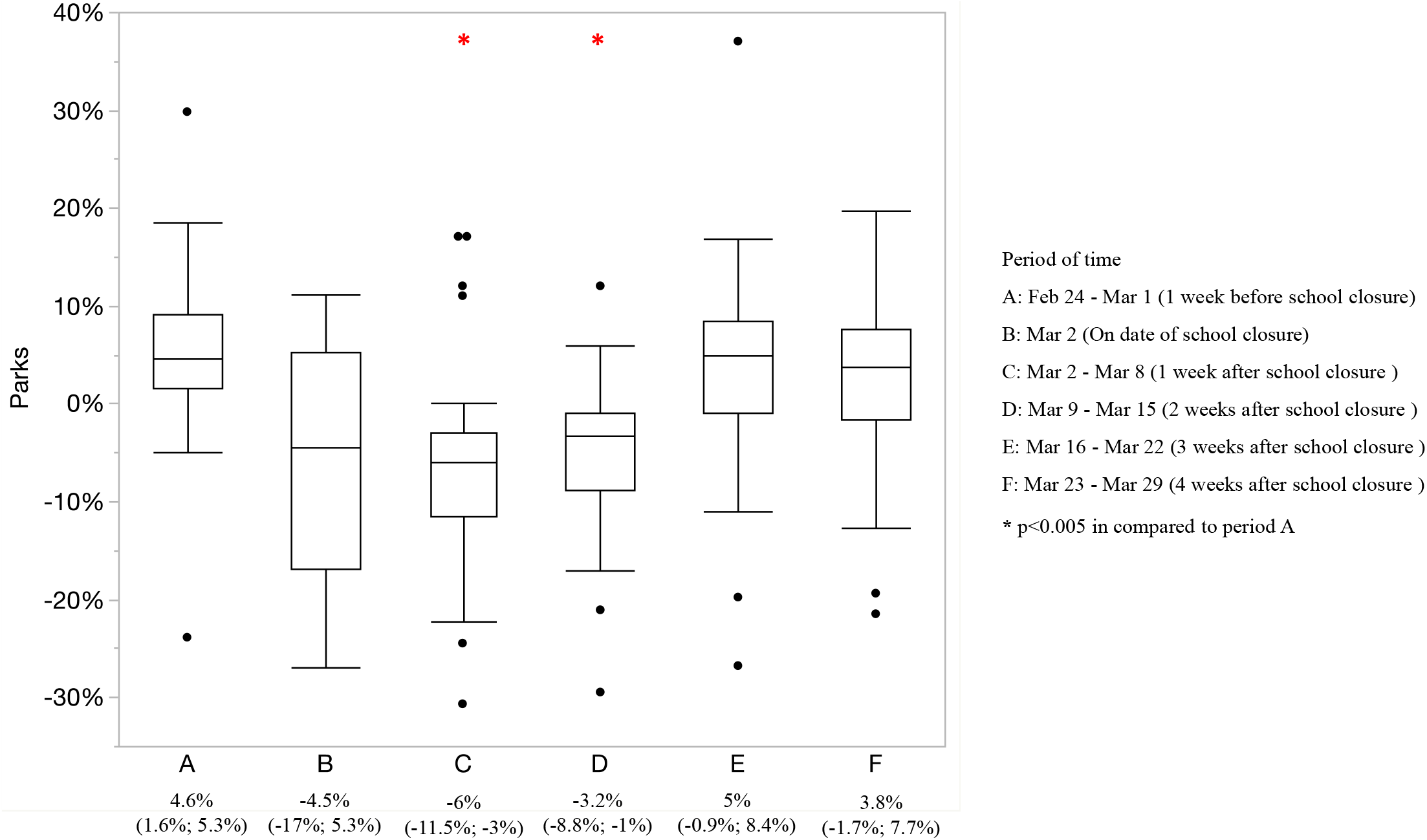
Mobility trend in parks of Japanese citizens from February 24^th^ to March 29^th^.

As expected, the percentage of transit stations experienced a decrease of nearly 10% over five weeks. The median (interquartile) mobility percentage of this category at period C, D, E, and F significantly reduced compared to period A (−21%(−25%;-19%); -20%(−24%;-17%); -17.7%(−22%;-14%), and -18.5%(−22%;-15%) vs. -9.8%(−13.5%;-6.9%)) (Figure 4).

**Figure 4:**
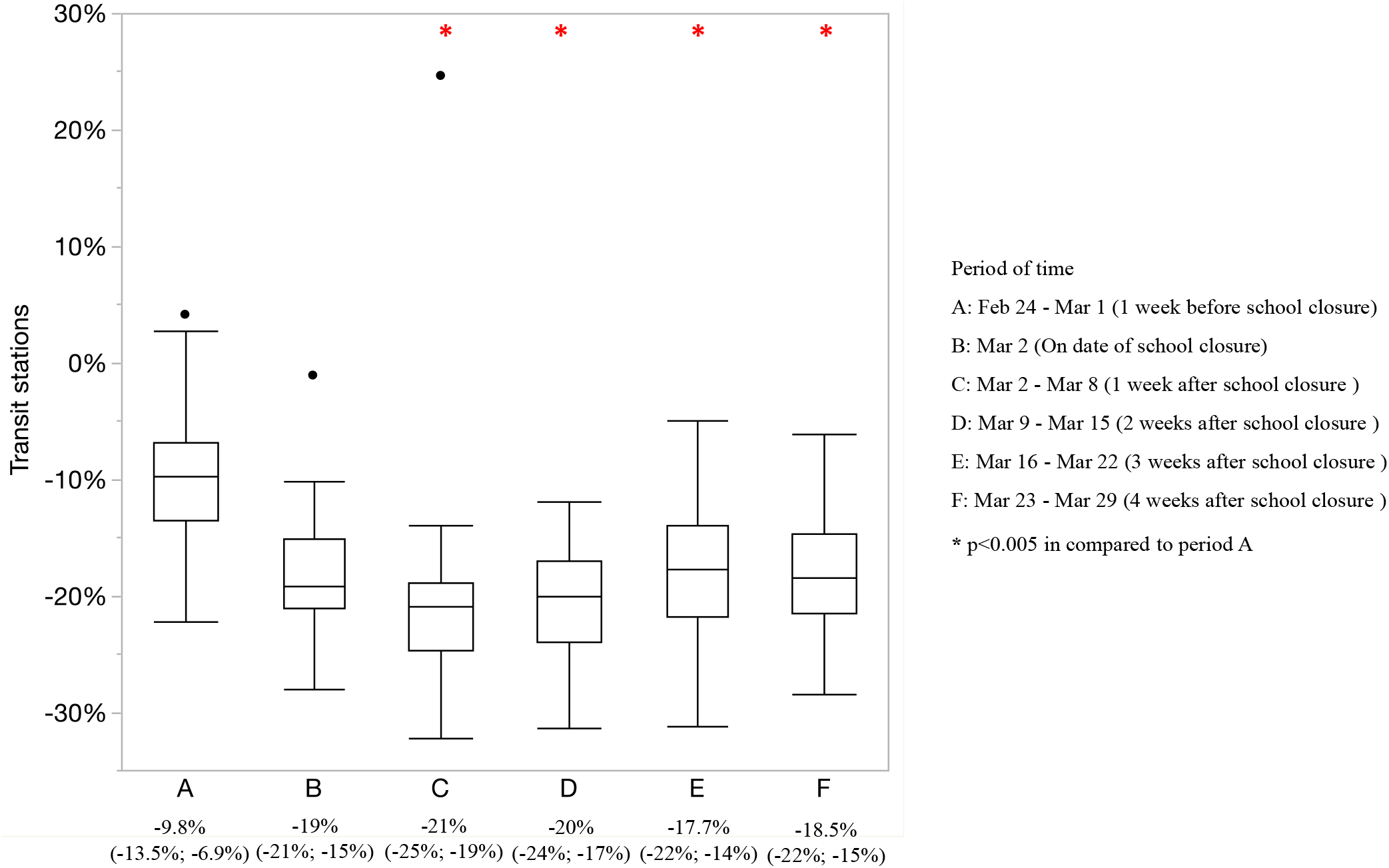
Mobility trend in transit stations of Japanese citizens from February 24^th^ to March 29^th^.

In contrast, the median (interquartile) mobility of Japanese residents for the workplace rose nearly 5% in period B, C, and D. There was a significant increment of mobility in workplace at one week and two weeks after school closure compared to one week before school closure (−2.9%(−4%;-1.5%); -3%(−4.5%;-2.1%) vs. -7.7%(−13.5%;-4.4%)). Afterwards, the workplace mobility decreased in the next two weeks and there was no significant difference in the mobility percentage of the workplace when comparing period A to period E and F (Figure 5).

**Figure 5:**
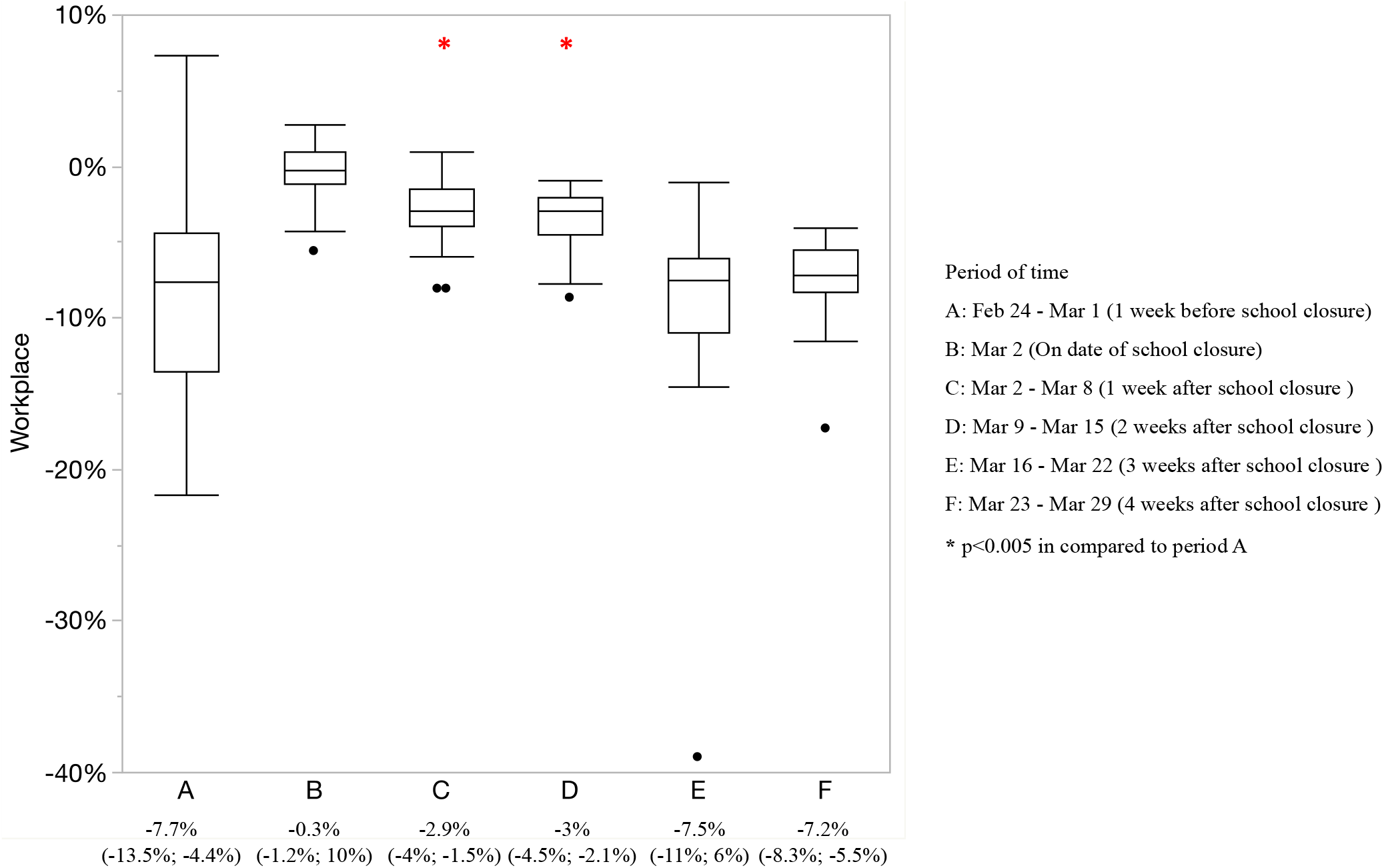
Mobility trend in the workplace of Japanese citizens from February 24^th^ to March 29^th^.

Concerning the mobility of residential, a trend of augmentation was exhibited since the date of school closure. The median (interquartile) mobility percentage of residential significantly increased in period C, D, E, and F compared to period A (3%(2.8%;4%); 3.1%(2.5%;3.9%); 3.4%(2.8%;4%), and 3.6%(3%;4.5%) vs. 2.7%(2.5%;3.1%) (Figure 6).

**Figure 6:**
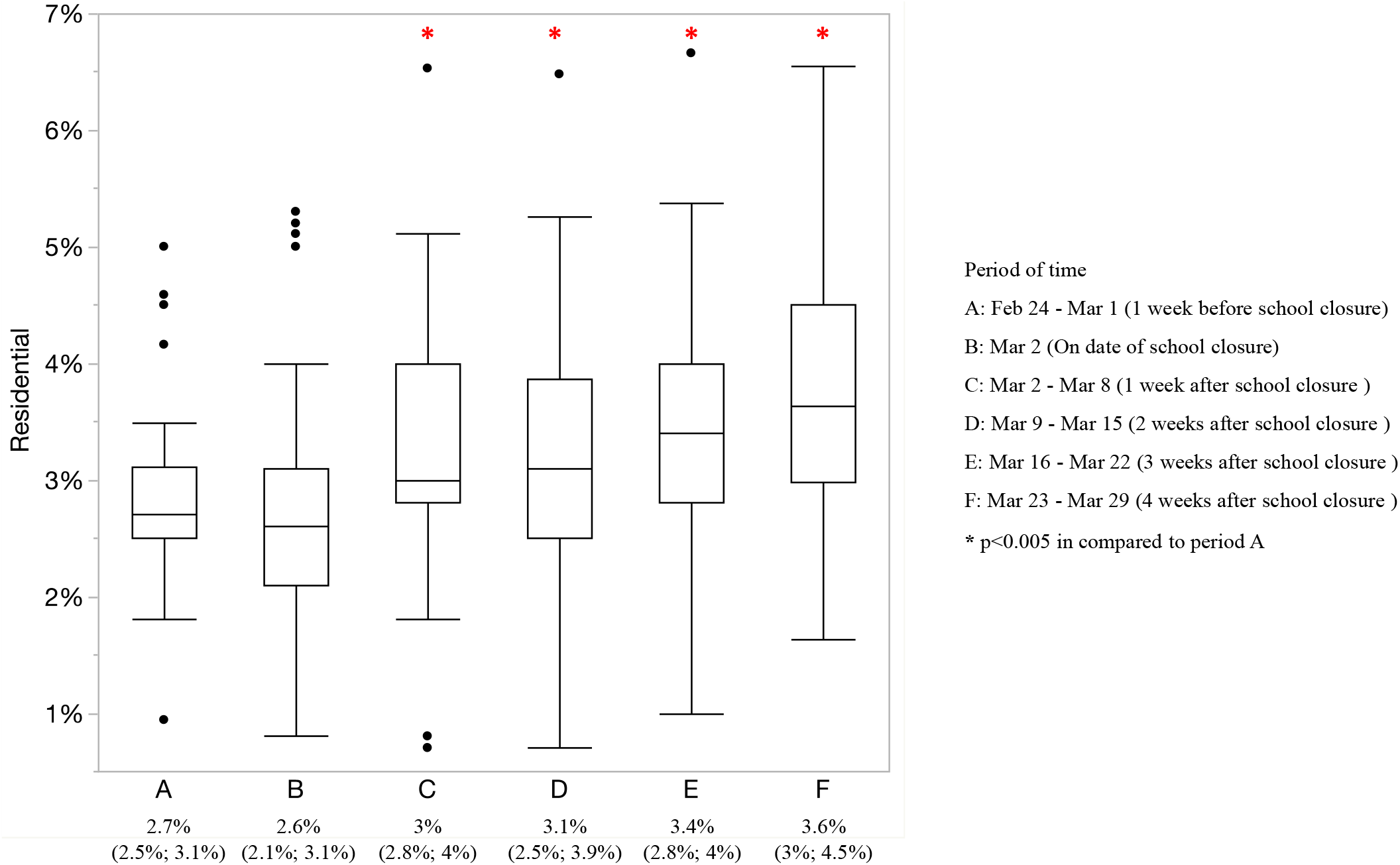
Mobility trend in residential of Japanese citizens from February 24^th^ to March 29^th^.

## Discussion

Governments all over the world have taken several actions in order to prevent further spread of COVID-19 that started on December 31^st^, 2019^1^. During the pandemic, due to the unavailability of authorized vaccines and effective antiviral therapies, school closure was represented as one of the main control measures to mitigate the circulation of COVID-19 in many countries in the world even though its efficacy is still debatable^9,13^. Based on the evidence retrieved from the experience of the seasonal and pandemic influenza outbreaks^6^, the possibility of transmitting the disease via direct contacts^14^, and a universal consensus of COVID-19 control measures^15^, school closure in the era of COVID-19 is expected to limit the infection rate, especially within the vulnerable population such as students. School closure was applied in Japan from early March to June^10^ in compliance with a government advisory and the measure consequently changed the daily activities of Japanese citizens. By using the COVID-19 Community Mobility Report from Google to obtain the mobility data of 47 prefectures in Japan, our study determined a significant difference in mobility of Japanese citizens regarding six basic indicators, including retail and recreation, grocery and pharmacy, parks, transit stations, workplace, and residential at two periods of time which are before and after school closure.

At first, a significant reduction in the mobility of retail and recreation, grocery and pharmacy, and parks were detected at one week after school closure. Even though there was a trend of recovery, the rate of Japanese citizens involving in these two categories after application of school closure remarkably declined compared to the prior period. The worse situation of the outbreak as well as the implementation of school closure undeniably imposed a noticeable influence concerning daily activities of Japanese citizens. The volume of people going to routine locations (supermarkets, convenience stores, etc.), closed spaces (restaurants, coffee shops, etc.), and public areas (parks, beaches, etc.) was deteriorated. School closure served as a warning sign that indicated the severity of the outbreak. Therefore, citizens became more cautious in their mobility and avoided going out to reduce the risk of catching COVID-19. Moreover, the Japanese government emphasized the importance of social distancing along with the announcement of school closure. The decrease in mobility of retail and recreation, grocery and pharmacy, and parks demonstrated the compliance, responsibility, and awareness of Japanese citizens in respecting the recommendations of the government.

Regarding public transportation, starting from March 2^nd^, a significant reduction in the volume of Japanese citizens using buses, railways, and subways was identified. Regardless of the normal operation of transit stations in Japan, this decline was therefore due to the absence of students who account for a large volume of daily customers. Additionally, recommendations from the government to limit the translocation and the travel to other prefectures also contributed to this mobility reduction. Furthermore, the mobility of Japanese citizens in workplaces decreased from the moment of two weeks after school closure. The requirement of working from home was adopted by the government and private sector companies to ensure the safety of employees and mitigate the spread of the virus by limiting contacts among employees. Besides the fact that employees were encouraged to work at home via online platforms and therefore reducing direct contacts, school closure also imposed parents to stay at home to take care of their children. Moreover, due to COVID-19, nearly 25 million jobs were lost, resulting in an increase of worldwide unemployment rate from 4.936% to 5.644%^16^. Along with the transition to telecommuting, the increase in the rate of unemployment during the pandemic contributed to the reduction of mobility in workplaces.

Finally, as expected, the results showed that the mobility trend in residential after school closure significantly increased compared to the prior period. Since students no longer went to schools, parents might take some days off or even quit jobs in order to take care of their children. Hence, school closure not only increased mobility in residential of students and educators but also those who did not work at schools. In addition, recommendations from the government to restrict the translocation encouraged Japanese citizens to stay at home. Also, in an effort to limit social contacts, telecommuting began replacing the conventional working strategies, thus leading to an increase in the number of people staying at home.

Besides, we acknowledge that our study has several limitations. Since Google COVID-19 Community Mobility Reports only recorded the location history of people who had a Google account and enabled the setting for location history, data of our study cannot represent the mobility trend of the whole Japanese population. Additionally, evaluation of the influence of school closure was restricted within four weeks since its application. Thus, this physical distancing measure requires a monthly assessment to identify its long-term potency.

In conclusion, the impacts of school closure could diversely vary among countries with different population structures and various cultural identities. By using the COVID-19 Community Mobility Report from Google, our study quantified the short-term effects of school closure on the mobility of Japanese citizens for retail and recreation, grocery and pharmacy, parks, transit stations, workplace, and residential and determined a significant change in the mobility trend of Japanese citizens before and after school closure. In Japan, even though the policies and the control measures of the government to manage the outbreak just stopped at the level of strong recommendation but not compulsion, these data reflected the responsibility and the awareness of Japanese citizens to a certain degree. While the efficacy of school closure is still debatable, its capability to control the pandemic cannot be ignored. Closing schools had reduced the social interactions among people and had eventually mitigated seasonal and pandemic influenza^7^. Therefore, it is highly recommended to close schools nationwide during the COVID-19 pandemic because school closure is an effective strategy to reduce the mobility of people, thus preventing human-to-human transmission.

## Data Availability

The data that support the findings of this study are available from the corresponding author, NTH, upon reasonable request.

## Acknowledgements

None.

## Authors’ contributions

NHN analyzed data and drafted the manuscript. APNN extracted data and drafted the manuscript. BTDL, AGG, ASMAA, AS, and AGP extracted data. VDK provided guidance on using the software to extract data. NTH organized the work and verified all data and the manuscript. All authors read and approved the final manuscript.

